# Clinicopathological Evaluation of Dry eyes and Ocular surface in Newly diagnosed patients of Hyperthyroidism and Hypothyroidism and its Comparison to Healthy Subjects

**DOI:** 10.1101/2020.12.15.20248298

**Authors:** Nidhi Paharia, Nikhil Agrawal, Shruti Agrawal

**Affiliations:** AIIMS Bhatinda; Aiims rishikesh

**Keywords:** Hyperthyroidism, Hypothyroidism, Impression Cytology, Ocular surface Inflammation

## Abstract

**Purpose:** To evaluate tear function test, corneal surface disorder in recently diagnosed hyperthyroid and hypothyroid patients and to compare results with healthy subjects.

**Setting/Venue:** Mahatma Gandhi Medical College, Jaipur, Rajasthan, India

**Methods:** Cross sectional analytical study. 30 recently diagnosed (within 3 months of diagnosis) patients of hyperthyroidism and hypothyroidism each and 30 healthy age matched controls were included. All patients and controls underwent assessment of proptosis, Palpebral fissure-height (PFH), tear function test (OSDI questionnaire, Schirmer’s test and Tear Break-up-time (TBUT) and Impression Cytology for ocular surface assessment. These parameters were compared between cases and controls. Standard statistical analysis was used.

**Results:** Mean Proptosis and PFH showed no significant difference among groups (P value =0.071). Mean value of OSDI was 42.33+22.67, 41.15+16.03 and 29.33+6.84 in hyperthyroid, hypothyroid & Controls respectively, the difference being statistically significant (p=0.001).

Mean TBUT was 7.13+3.28 sec, 6.38+2.46 sec, and 11.15+2.39 sec in hyperthyroidism, hypothyroidism and controls respectively (p=0.001). Mean value of Schirmer tear test was 12.93+5.81mm, 13.30+4.44mm, and 17.55+7.35mm in hyperthyroidism, Hypothyroidism and controls respectively (p=0.001). 80% of patients in hyperthyroidism group had grade 2-3 squamous metaplasia as compared to 70% in hypothyroidism patients and 24.4% in controls, signifying ocular surface damage (p<0.05).

**Conclusions:** Mean PFH and proptosis did not differ between three groups. However, increased OSDI score, decreased Schirmer’s test value, decreased TBUT and grade 2-3 squamous metaplasia in patients of thyroid dysfunction suggests presence of dry eyes. Despite no clinically visible signs of thyroid ophthalmopathy, there is ocular surface damage right from the early stages of thyroid dysfunction. This is attributable to evaporative mechanism as well as ocular surface inflammation and hyperosmolarity of tear film.

## Introduction

Thyroid disorders, both hyperthyroidism and hypothyroidism, are commonly prevalent worldwide, often affecting females more than males^1,2^. Thyroid disorders are often associated with ophthalmic manifestations, commonly referred to as thyroid eye disease (TED) or thyroid-associated ophthalmopathy (TAO). The prevalence of TED in hypothyroidism is 0.2-8.6%, whereas in hyperthyroid patients it is as high as 50%.^3-5^ The approximate time from the onset of thyroid disease to the clinical manifestation of TED is 18 months.^3,6^ Dry eyes is one of the most common manifestation in patients with TED, with a reported incidence varying between 23-96%.^2,3^ It is a multifactorial disease caused by increased surface evaporation of the tear film due to mechanical factors (lid retraction, increased palpebral fissure height, and exophthalmos), altered tear production due to immunological factors (ocular surface damage, alteration in tear film composition, and damage to the lacrimal glands.^6,7^ According to the literature, 37% of patients with Hashimoto’s disease and 72% of patients with Grave’s disease have reported symptoms of dry eyes.^8,9^

Conjunctival impression cytology, which was first introduced by Egbert et al in 1977, is a non-invasive, simple technique that provides information about morphology of the superficial layers of the ocular surface. ^10-12^ In fact, it has been reported that ocular surface inflammation resulting in squamous metaplasia of the conjunctival epithelium, which can be demonstrated on conjunctival impression cytology, may be the earliest clinical sign in Graves’ disease before the onset of clinically evident TAO.^13-16^ Although the association of dry eyes with hyperthyroidism is well documented^13,14,15^, there is a paucity of literature documenting the presence of dry eyes in hypothyroid patients.^9,16,^ The present study aims at studying the presence of dry eyes with associated ocular surface inflammation and consequent squamous metaplasia of the conjunctival epithelium in recently diagnosed patients of hypothyroidism and hyperthyroidism.

## Material and methods

This single point, observational, cross-sectional study was conducted in the department of ophthalmology at a tertiary eye centre in Western India. Prior approval for the study was obtained from the Institutional Ethics Committee and informed consent taken from all subjects before their enrollment for the study. The study was conducted following the tenets of the declaration of Helinski. Patients who were diagnosed with hypothyroidism or hyperthyroidism within 3 months from the date of examination were selected randomly from the endocrinology department of the same hospital. Cases were defined as patients with primary hypo/hyperthyroidism diagnosed for the first time based on a combination of laboratory investigations and clinical diagnosis by an endocrinologist. The cases were further classified as group 1 (30 patients) – patients having hypothyroidism, or group 2 (30 patients) – those having hyperthyroidism. Patients with a history of intake of any form of topical ocular medications for pre-existing eye diseases, prior ocular trauma or surgery, glaucoma, uveitis, contact lens wear, any known systemic autoimmune disorder or disorders affecting ocular motility (eg. myasthenia gravis, Lambert Eaton syndrome), diabetes mellitus and hypertension, were excluded from the study.

Age and gender-matched healthy controls (30 in number) were randomly chosen from among patients attending the ophthalmology clinic for refraction check without presenting symptoms of dry eyes and without any ocular comorbidity. This group was labelled as group 3. Only one eye of each patient was randomly selected for inclusion in the study in order to avoid selection bias.

### Protocol for ocular evaluation

All the cases and controls underwent a complete ophthalmic examination of both eyes including visual acuity testing, pupillary reaction, slit-lamp examination, applanation intraocular pressure measurement, fundus examination and ocular motility assessment. Exophthalmometry was performed for both eyes of each patient for assessment of proptosis, using the Hertel’s exophthalmometer. Palpebral fissure height (PFH), presence of lid retraction and lid lag were also documented for each eye. All patients underwent the following tear function tests: ocular surface disease index (OSDI), Schirmer tear test and tear film break-up time (TBUT). The symptoms of dry eye were assessed according to the OSDI questionnaire, based on a one-week recall period. OSDI subscale scores can range from 0 to 100, with higher scores indicating more problems or symptoms. The index demonstrates sensitivity and specificity in distinguishing between normal subjects and patients with dry eye disease. A wetting of less than 10mm was taken as abnormal on the Schirmer I tear test (performed without topical anesthesia). To measure TBUT, a fluorescein sodium strip moistened with a drop of non-preserved saline solution was applied to the inferior palpebral conjunctiva in each eye of the patients and the control subjects. The pre-corneal tear film was examined with a bio-microscope, and the elapsed time before the initial breakup, rupture of the tear film or formations of tiny dry spots were recorded. A TBUT below 10 seconds was taken as abnormal.

### Methodology for conjunctival impression cytology

Conjunctival impression cytology was performed in all patients to study the morphologic characteristics of the ocular surface. Samples were collected from the temporal interpalpebral bulbar conjunctiva. Cellulose acetate filter was used to obtain samples. The filter paper was cut asymmetrically to form a quadrilateral with 4 mm in height, 5 and 6 mm in length for temporal bulbar conjunctiva. The filter paper was placed keeping the matt surface on the conjunctival side, 3mm behind the limbus and gentle pressure was applied to provide better adherence to the conjunctival tissue. The paper was removed with a non-toothed forceps and fixated in a solution of 1:1:20 glacial acetic acid, 37% formaldehyde and 70% ethyl alcohol, and stained with periodic acid Schiff (PAS) and hematoxylin according to the protocol described by Tseng (1985)^11^. After mounting the paper, it was examined under the light microscope, and the cytologic changes were graded according to Nelson’s grading system as follows(Figure1 and Figure 2):

Grade 0: Small and round epithelial cells with a Nucleo-cytoplasmic ratio of 1:2 and eosinophilic cytoplasm; abundant, plump, oval goblet cells with intensely PAS-positive cytoplasm.

Grade 1: Slightly larger and more polygonal epithelial cells with Nucleo-cytoplasmic ratio 1:3 and eosinophilic cytoplasm. There is a decrease in goblet cell number.

Grade 2: Larger and polygonal, occasionally multi nucleated epithelial cells with variable staining cytoplasm and a Nucleo-cytoplasmic ratio 1:4-1:5. Smaller and less intensely PAS-positive goblet cells with poorly defined cellular borders and markedly decreased in number.

Grade 3: Large polygonal epithelial cells with basophilic cytoplasm. Small, pyknotic and in many cells completely absent nuclei with a nucleo-cytoplasmic ratio greater than 1:6. Goblet cells absent.

**Figure 1:**
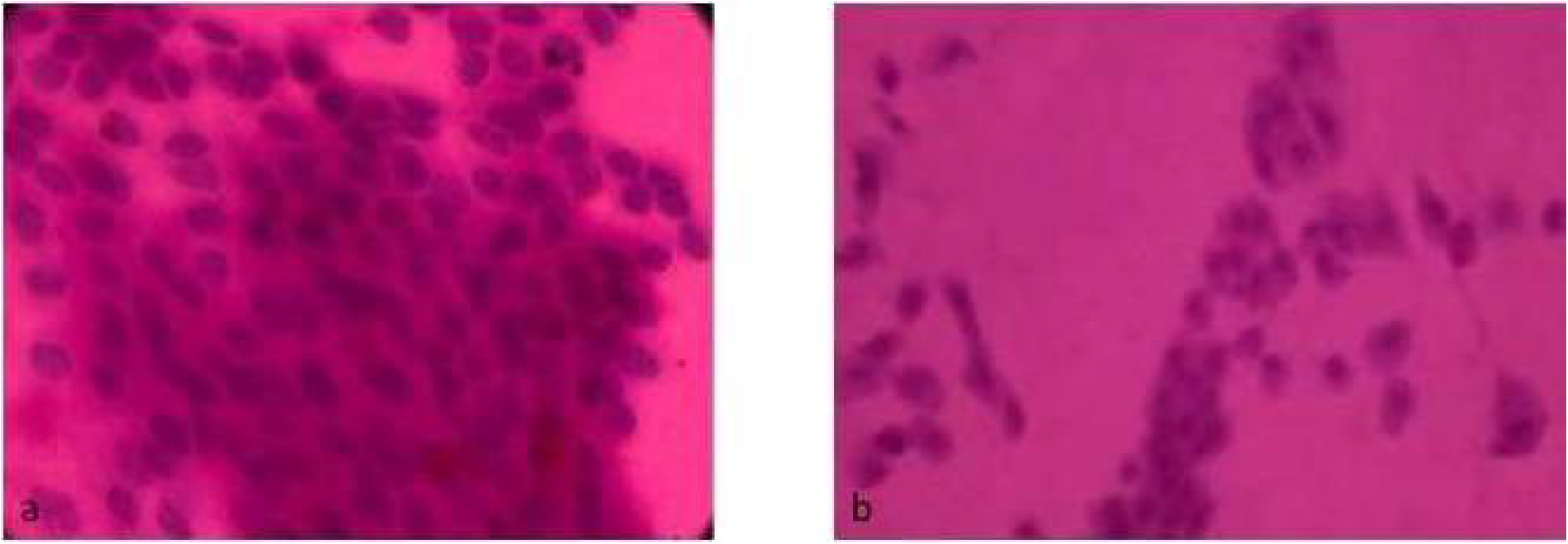
**a.Grade 0**: **Epithelial Cells**- Small & round, Nucleus: Cytoplasmic (N/C) ratio1:2 **Goblet Cells**: Abundant, plump, oval with PAS positive cytoplasm b. **Grade 1: Epithelial Cell**s-slightly larger and more polygonal with N/C ratio 1:3. **Goblet Cell** : Decrease in cell number.

**Figure 2:**
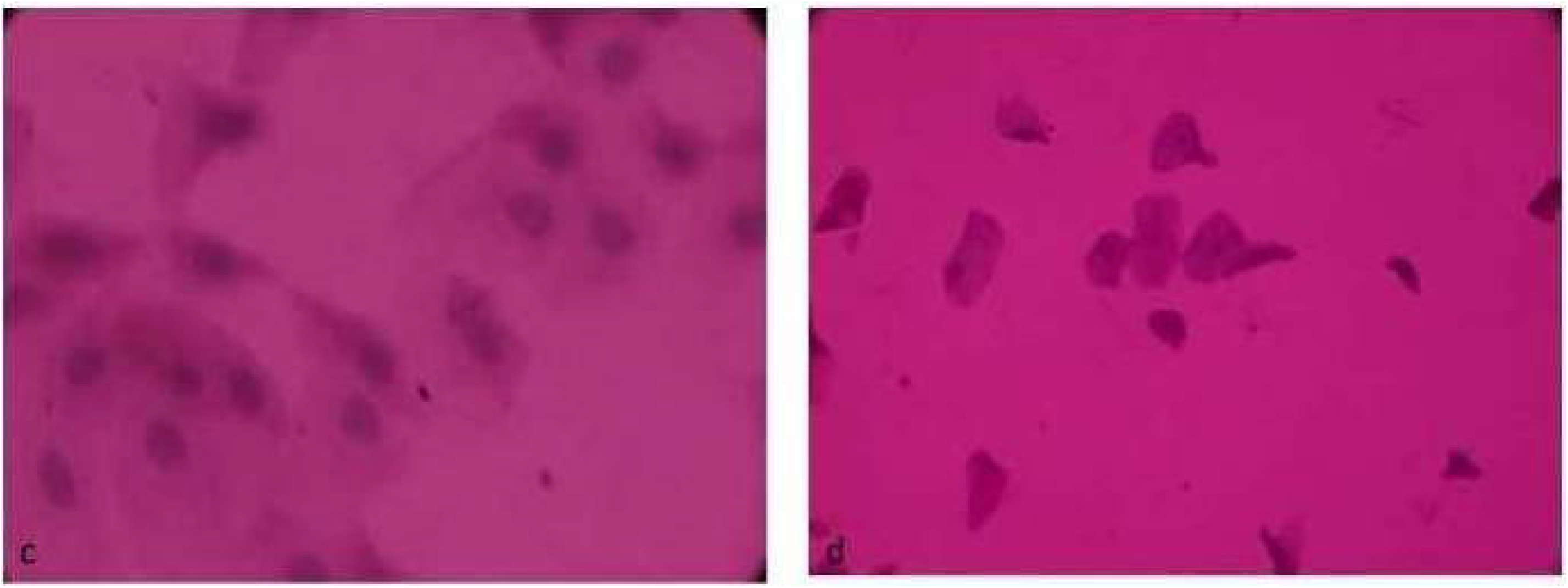
**a. Grade 2: Epithelial Cell**: Larger and polygonal, with N/C **Ratio 1:4-1:5** with poorly defined cellular border **Goblet cell**: Marked decrease in number. b. **Grade 3: Epithelial cell:** Large & polygonal with Small, **Pyknotic** & sometimes Absent nuclei. N/C ratio greater than 1:6. Goblet cells are absent

### Outcome Measures

The main outcome measures were to compare the tear film indices (OSDI scores, Schirmer test, TBUT) and impression cytology grading between cases (Groups I and II) and controls (Group III).

### Statistical analysis

The data including demographic profile, tear film indices and conjunctival impression cytology grading was entered in Microsoft Excel^®^. Statistical analysis was performed using the SPSS software (IBM, USA) version 19.0. Analysis of Variance (ANOVA) test was used to compare differences in mean OSDI, TBUT, and Schirmer tear test between groups. The impression cytology findings were compared by Chi-square test. Differences were considered statistically significant at a p value of <0.005.

## Results

The mean age of patients in Group I was 46.3 + 7.5 years, Group II was 47.3 + 8.5 years and Group III was 44.6 + 6.4 years (Mean±SD, p=0.618). There was an obvious female predominance in the patient distribution in the cases as well as controls with the male to female ratio of 1:3, 1:2 and 1:2 in group I, II and III respectively (p=0.386). The dry eye symptoms were significantly higher in cases i.e. dysthyroid patient as compared to the healthy controls. Findings of the ocular examination are presented in Table 1. Both, Groups I and II had a higher palpebral fissure height as compared to Group III, although the difference was not statistically significant (p = 0.071). Similarly, exophthalmometry did not reveal statistically significant differences between the three groups. Ocular surface and dry eye assessment findings are shown in Table 2. Groups I and II had a significantly greater number of patients with dry eye disease as compared to the healthy controls with the OSDI being higher and the TBUT and Shirmer’s test being significantly lower in the dysthyroid patients as compared to the controls (p<0.05).

**Table 1:** Comparison of ocular measurements between the Groups within the study

| | Group I<br>(Hypothyroidism,<br>n=30)<br>(Mean± SD) | Group II<br>(Hyperthyroidism,<br>n=30)<br>(Mean± SD) | Group III<br>(Control,<br>n=30)<br>(Mean± SD) | P Value# |
| --- | --- | --- | --- | --- |
| Palpebral<br>fissure Height,<br>(mm) | 12.10 ± 1.22 | 12.30 ± 1.26 | 11.73 ± 0.94 | 0.115 |
| Proptosis<br>assessment<br>(mm) | 20 ± 1.8 | 20.30 ± 1.40 | 19.9 ± 1.40 | 0.582 |

- \*SD = standard deviation - #Pvalue of $<0.05$ considered significant, ANOVA test used

**Table 2:** Comparison of Ocular surface and Dry Eye Disease Assessment between the Groups

| | Group I<br>(Hypothyroidism, n=30)<br>(mean ± SD) | Group II<br>(Hyperthyroidism, n=30)<br>(mean ± SD) | Group III<br>(Control, n=30)<br>(mean ± SD) | P Value# |
| --- | --- | --- | --- | --- |
| Ocular Surface Disease Index (OSDI) | 41.15 ± 16.03 | 42.30 ± 22.67 | 29.23 ± 6.84 | 0.001 |
| Tear film Breakup Time (TBUT, seconds) | 6.38 ± 2.46 | 7.13 ± 3.28 | 11.15 ± 2.39 | 0.001 |
| Schirmer's Test (wetting of strip, mm) | 13.30 ± 4.44 | 12.93 ± 5.81 | 17.55 ± 7.35 | 0.002 |

- \*SD = standard deviation - #P value of < 0.05 considered significant, ANOVA test used

As shown in table 3, the assessment of the conjunctival impression cytology specimen collected from temporal inter-palpebral conjunctiva showed 70% and 80% grade 2-3 squamous metaplasia in hypothyroid and hyperthyroid patients respectively as compared to 26.6% in healthy controls (p=0.001).

**Table 3.** Incidence of squamous metaplasia on conjunctival impression cytology – comparison between the Groups

|  | Group 1<br>(Hypo) | Group 2<br>(Hyper) | Group 3<br>(Control) | P Value |
| --- | --- | --- | --- | --- |
| Grade 0-1 | 9(30%) | 6(20%) | 22(73.3%) | <0.001 |
| Grade 2-3 | 21(70%) | 24(80%) | 8(26.6%) | <0.001 |

#P value of < 0.05 considered significant, chi-square test used

## Discussion

In dysthyroid ophthalmopathy, proptosis and increased PFH have been conventionally considered to be the major contributing factor in the etiology of dry eyes by causing increased evaporation of tear film due to impaired blinking.^17^ Furthermore, it has been postulated that degree of tear film evaporation is proportional to the ocular surface area exposed.^18^ However, in the present study, a majority of the patients with both hyperthyroidism and hypothyroidism showed deranged tear function tests, and presence of squamous metaplasia on impression cytology even without the presence of proptosis and increased PFH. This suggests the role of an alternative mechanism in the pathogenesis of dry eyes in cases of thyroid dysfunction.

Ophthalmic evaluation of subjects showed that OSDI was in severely impaired range in both hypothyroid and hyperthyroid patients, which was in concordance with the findings of Gurdal et al and Turkyilmaz et al. ^15,19^ Despite normal PFH, OSDI score, in comparison to normal healthy controls, was in mild to moderate dry eye zone, further correlating presence of high score in an asymptomatic individual.

Schirmer’s score was also found to be reduced in patients with thyroid dysfunction as compared to healthy subjects in this study. This finding was congruous with the results reported by Eckstein et al in their study. They identified presence of TSH receptors not only on the lacrimal gland but also on corneal and conjunctival epithelium which acted as target for autoantibodies in thyroid disease resulting in lacrimal gland impairment and subsequent attenuation in tear secretion.^20^ Villani et al concluded that a reduced Schirmer’s score in patients with Grave’s ophthalmopathy with normal PFH was due to diminished corneal sensitivity due to ocular surface inflammation explaining the occult character of dry eyes in these patients.^21^

The Tear break up time (TBUT) was significantly decreased in hyperthyroid and hypothyroid patients in the present study suggesting an unstable tear film in comparison to the control subjects. According to U.S. National Eye Institute (NEI) Committee on dry eyes, the instability of tear film could be caused by hyperosmolarity of tear film resulting from a decrease in tear secretion due to a lacrimal gland disease or from an increase in tear film evaporation.^17^ On the other hand, Kan et al discovered the role of inflammation in addition to tear film hyperosmolarity in the development of dry eyes in TAO.^16^

Impression cytology is the standard technique to study the effect of chronic inflammation in the form of squamous metaplasia and loss of goblet cells in ocular surface disorder. In the current study, a greater proportion of the patients with hyperthyroidism as well as hypothyroidism had grade 2-3 squamous metaplasia in comparison to the healthy controls. Persistent hyperosmolarity of the tear film and ocular surface inflammation may cause pathological changes in corneal epithelium, such as increased desquamation, blunting and loss of microplicae, disruptions of the cell membrane and cellular swelling with decreased goblet cell density. These cascades of events lead to squamous metaplasia, reduced goblet cell density and larger polygonal epithelial cells with a decreased nucleo-cytoplasmic ratio which can be picked up by a meticulously performed impression cytology technique.^22^ Besides, presence of lymphocytes in conjunctival impression cytology as observed in patients of thyroid dysfunction as compared to controls further strengthen the role of ocular surface inflammation.^19^

Hoffman et al demonstrated that deficiency of thyroid hormone (TH) in hypothyroid patients predisposes to ocular surface structural changes.^23^ The possible pathophysiology could be the increase in oxidative stress caused due to alteration in oxidative metabolism by reduced levels of TH which normally regulates the function of lacrimal glands, cornea & conjunctiva through TSH receptors. Hence, a chronic low thyroxine level has an impact on both tear secretion and ocular surface inflammation with resultant alternation in TBUT and Schirmer’s test as confirmed by impression cytology findings.^24-26^

The etiopathogenesis of dry eye-related with TED has been investigated in several recent studies. According to Luo et al, tear film hyperosmolarity has been shown to induce dry eye syndrome through inflammatory cytokines such as interleukin-1 (IL-1), TNF-alpha, MMP-9 which activates mitogen active protein kinase signal (MAPK) pathway leading to ocular surface damage and dry eyes.^27^ (Figure 3,4) Iskeleli et al postulated that disruptions of the normal composition, quantity, and physiology of the ocular tear film can cause a vicious cycle of increased tear film evaporation and subsequent dry eye symptoms.^28^ The present study however emphasises upon the role of ocular surface inflammation as a causative factor for dry eyes because of the presence of an unstable tear film even in absence of proptosis and increased PFH in patients of thyroid disorder.

**Figure 3:**
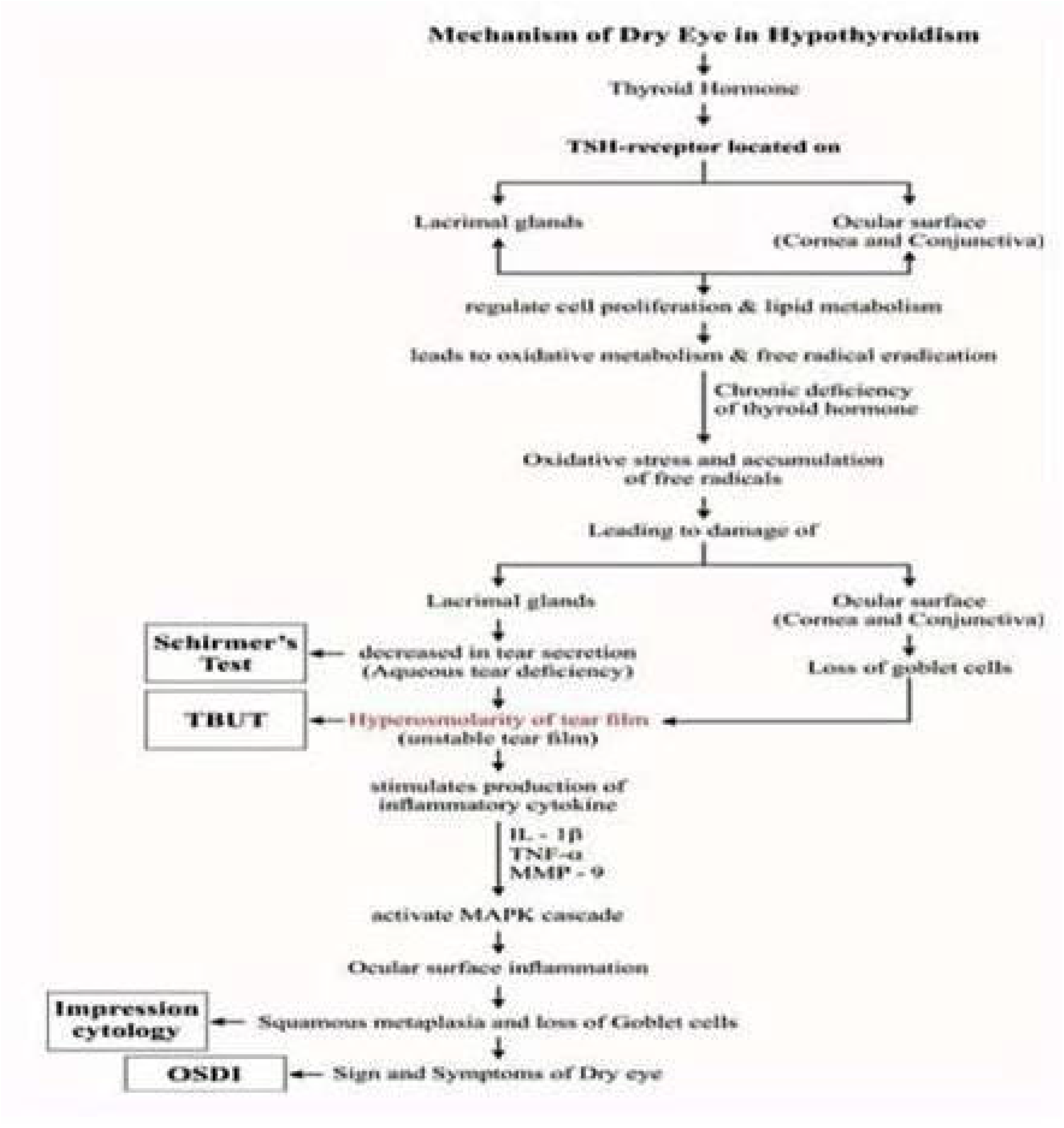
Mechanism of Dry eyes in Hypothyroidism

**Figure 4:**
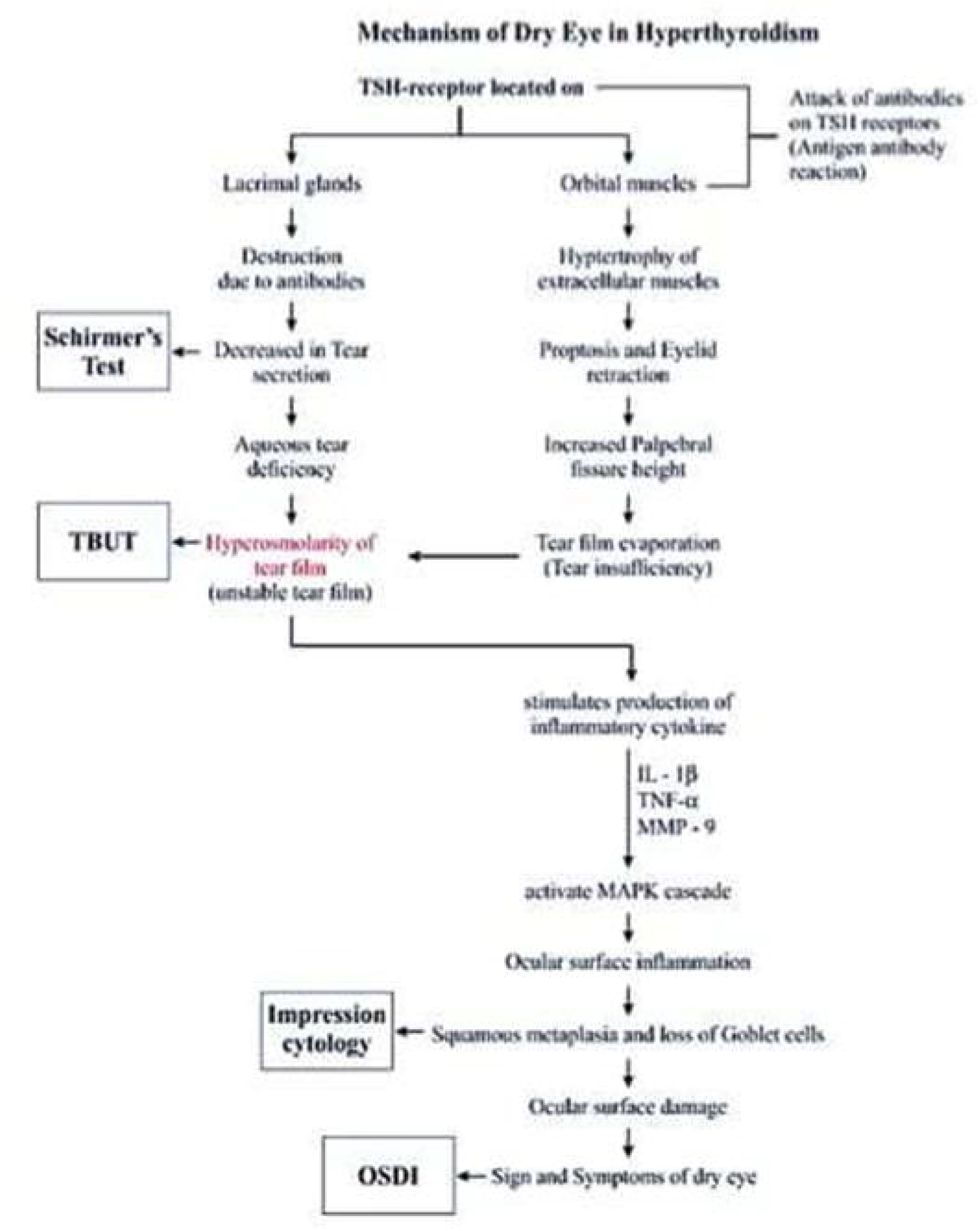
Mechanism of dry eyes in Hyperthyroidism

In view of our findings, we recommend that the thyroid profile must be performed as a routine investigation in patients diagnosed with dry eye, especially among women of 30-50 years. Proactive ocular surface & dry eye evaluation should be performed in patients diagnosed with hyperthyroidism & hypothyroidism. In a high volume OPD, TBUT should be performed in all patients of thyroid dysfunction. To summarise, an ophthalmologic referral is mandatory in any patient diagnosed with thyroid dysfunction.

The limitations of the study include a small sample size apart from lack of objective evaluation of meibomian glands, non-availability of inflammatory markers in impression cytology and non-measurement of hyperosmolarity of the tear film.

## Conclusion

Significant conjunctival squamous metaplasia is present in dysthyroid patients in contrast to the euthyroid patients inspite of no significant difference in the mean PFH and proptosis between the two groups. We suggest that the deranged tear function in patients of thyroid dysfunction is attributed to ocular surface inflammation due to unstable tear film (hyperosmolarity) rather than tear film evaporation.

## Data Availability

All the data mentioned in the manuscript is available in the manuscript attached.

## Notes

### Competing Interest Statement

The authors have declared no competing interest.

### Funding Statement

None

### Author Declarations

Institutional Ethics committee, Mahatma Ganghi Medical College, Jaipur

